# Booster of mRNA-1273 Strengthens SARS-CoV-2 Omicron Neutralization

**DOI:** 10.1101/2021.12.15.21267805

**Authors:** Nicole A. Doria-Rose, Xiaoying Shen, Stephen D Schmidt, Sijy O’Dell, Charlene McDanal, Wenhong Feng, Jin Tong, Amanda Eaton, Maha Maglinao, Haili Tang, Kelly E. Manning, Venkata-Viswanadh Edara, Lilin Lai, Madison Ellis, Kathryn Moore, Katharine Floyd, Stephanie L. Foster, Robert L. Atmar, Kirsten E. Lyke, Tongqing Zhou, Lingshu Wang, Yi Zhang, Martin R Gaudinski, Walker P Black, Ingelise Gordon, Mercy Guech, Julie E Ledgerwood, John N Misasi, Alicia Widge, Paul C. Roberts, John Beigel, Bette Korber, Rolando Pajon, John R. Mascola, Mehul S. Suthar, David C. Montefiori

## Abstract

The Omicron variant of SARS-CoV-2 is raising concerns because of its increased transmissibility and potential for reduced susceptibility to antibody neutralization. To assess the potential risk of this variant to existing vaccines, serum samples from mRNA-1273 vaccine recipients were tested for neutralizing activity against Omicron and compared to neutralization titers against D614G and Beta in live virus and pseudovirus assays. Omicron was 41-84-fold less sensitive to neutralization than D614G and 5.3-7.4-fold less sensitive than Beta when assayed with serum samples obtained 4 weeks after 2 standard inoculations with 100 µg mRNA-1273. A 50 µg boost increased Omicron neutralization titers and may substantially reduce the risk of symptomatic vaccine breakthrough infections.

## Text

Over the last two years, the COVID-19 pandemic has produced successive waves of SARS-CoV-2 variants of concern (VOCs) that outcompete earlier variants, escape therapeutic antibodies, and demonstrate partial resistance to neutralizing antibodies induced by natural infection and vaccination. The earliest variant carried a single spike mutation, D614G, providing a fitness advantage for transmission and rapidly replacing the ancestral virus as the dominant pandemic variant by May, 2020 (1). Importantly, D614G was dominant over the period of time when the Moderna mRNA-1273 vaccine demonstrated 94% efficacy in preventing symptomatic COVID-19 in the Coronavirus Efficacy (COVE) phase 3 trial (2, 3), making D614G spike protein a primary reference standard. D614G spike binding and neutralizing antibodies also correlated with vaccine efficacy in the COVE study (4), further strengthening the utility of this spike as a reference standard. Notably, D614G is one of the most neutralization-sensitive forms of the virus known to date (5). The Alpha variant approached dominant status by early 2021 and was soon surpassed by the Delta variant, which has dominated the pandemic since mid-2021. Delta was a modest neutralization escape variant, being 2-3 fold less susceptible than D614G to neutralization by mRNA-1273 vaccine-induced antibodies (5) and having little impact on mRNA-1273 vaccine efficacy (6). Other variants caused transient regional outbreaks but were unable to compete on a global scale. Among the latter, the Beta variant that circulated in South Africa in late 2020 to mid-2021 is among the least neutralization-sensitive variants characterized to date (5) but nonetheless remains susceptible to mRNA vaccines with only a modest reduction in efficacy, especially against serious illness and death (7).

The Omicron variant that was first identified in South Africa in November 2021 is raising concerns because of its increased transmissibility and potential for reduced susceptibility to antibody neutralization attributed to the presence of a large number of spike mutations, including 15 mutations in the receptor binding domain (RBD) that is the major target for neutralizing antibodies (8, 9). To assess the potential risk of this variant to existing vaccines, identical serum samples from participants who received two standard inoculations of mRNA-1273 (100 µg) in the COVE study were tested for neutralization of Omicron in lentivirus-based pseudovirus assay (4) in two independent laboratories: The Vaccine Research Center (VRC lab) in the National Institutes of Health (NIH), and Duke University Medical Center (Duke lab). A live virus focus-reduction neutralization (FRNT) assay was performed at Emory University (Emory lab) (10). One subset of samples from the COVE study was pre-selected as having high titers of neutralizing antibodies against D614G to enable detection of a wide range of titer reductions against Omicron. A second subset of COVE samples having moderate titers against D614G was pre-selected to avoid potential bias of high titer sera. The three laboratories assayed D614G and Beta as comparators.

The two pseudovirus-based assay laboratories also tested serum samples obtained from participants who received two inoculations of mRNA-1273 (100 µg) under Emergency Use Authorization (EUA) and later received a 50 µg boost of mRNA-1273, although here the two laboratories assayed samples from different subjects. The VRC lab tested serum samples from participants who received a third dose at 9 months after the second dose, all under EUA (VRC200 protocol). The Duke lab tested serum samples from a clinical study (NIAID heterologous boost study DMID 21-0012) designed to assess a vaccine boost at least 4 months after primary EUA-dosed vaccine recipients. The latter set specifically included the highest D614G responders.

The main purpose of this study was to gain an early understanding of the extent by which Omicron escapes vaccine-elicited neutralizing antibodies in comparison to D614G and Beta. The study was not designed to determine the full spectrum of vaccine-elicited neutralizing antibodies against Omicron. The Omicron spike in the pseudovirus and live virus used this study contained mutations A67V, Δ69-70, T95I, G142D, Δ143-145, Δ211, L212I, +214EPE, G339D, S371L, S373P, S375F, K417N, N440K, G446S, S477N, T478K, E484A, Q493R, G496S, Q498R, N501Y, Y505H, T547K, D614G, H655Y, N679K, P681H, N764K, D796Y, N856K, Q954H, N969K, L981F. The Beta variant in the pseudovirus and live virus contained spike mutations L18F, D80A, D215G, Δ242-244, R246I, K417N, E484K, N501Y, D614G and A701V. The D614G variant contained D614G as the only spike mutation in both viruses.

Compared to D614G, ID50 geometric mean titers (GMTs) after 2 doses of vaccine were 84-fold, 49-fold lower against Omicron in the VRC and Duke laboratories, respectively (Fig 1), and were 41-fold lower against Omicron in the Emory laboratory (Fig 2). This is a substantial decrease compared to the 13.6-fold, 9.2-fold, and 5.4-fold lower ID50 GMTs, respectively, against Beta. An approximate 12-fold improvement in Omicron neutralization was seen after the 50 µg boost such that the variant was now only 6.5- and 4.2-fold less neutralization-susceptible, respectively, than D614G in the pseudovirus assay (Fig 1). A post-boost improvement in neutralization also was seen for Beta (3.4- and 2.6-fold less susceptible, respectively, than D614G). Thus, the boost provided improvement in neutralization of both the Omicron and Beta variants. Minor differences in titer reductions between the two pseudovirus assay laboratories may be explained by sample sets that were not completely identical (see Fig 1 legend). Post-boost samples were not available to be assayed at the Emory laboratory.

**Figure 1.**
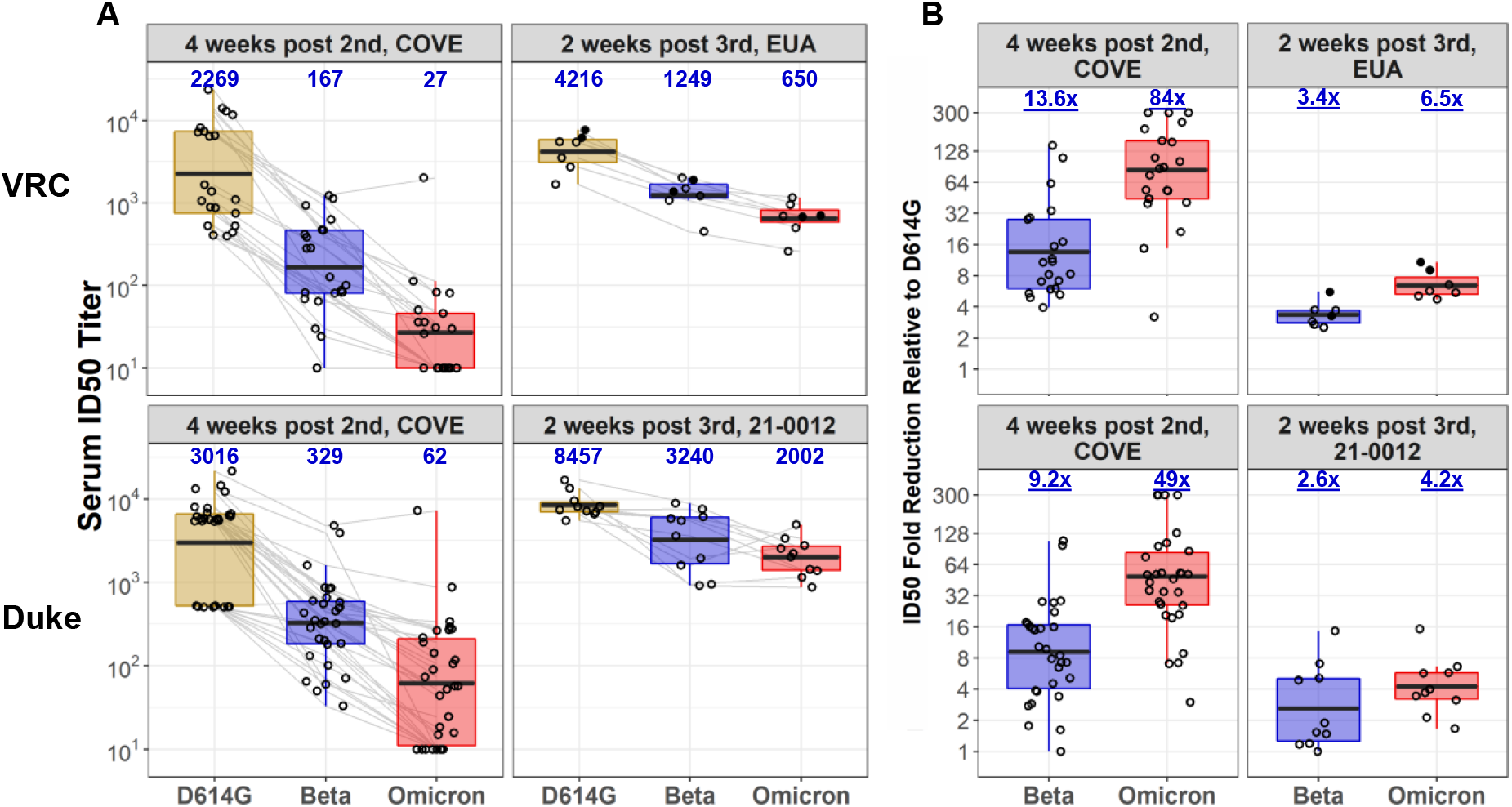
Neutralization of D614G, Beta and Omicron Pseudoviruses by Serum Samples Obtained from Moderna mRNA-1273 Vaccine Recipients. Serum samples obtained from 30 vaccine recipients 4 weeks after 2^nd^ inoculation with mRNA-1273 in the phase 3 COVE study (NCT04470427) were assayed against pseudoviruses bearing either the D614G, Beta or Omicron spike of SARS-CoV-2 in two independent laboratories (VRC: Vaccine Research Center, National Institutes of Health; Duke: Duke University Medical Center). Identical samples from the COVE study were assayed in both laboratories and were pre-selected for possessing either high (ID50 = 5,408 -22,573, n=20) or medium (ID50 = 506 - 526, n=10) neutralization titers against prototypic D614G. The Duke laboratory assayed 20 high titer and 10 medium titer samples. The VRC laboratory assayed 11/20 high titer samples and all 10 medium titer samples. Both laboratories also assayed serum samples from participants who received the primary series of two mRNA-1273 inoculations (100 µg) and a late boost (50 µg mRNA-1273). The late boost samples in the VRC laboratory were obtained 2 weeks after boosting in 7 participants who received the primary series under EUA (VRC200 protocol); the same 7 participants are shown in Fig 2. The late boost samples in the Duke laboratory were obtained 2 weeks after boosting in a phase 1/2 “Mix and Match” study (DMID 21-0012, NCT04889209) and were preselected for possessing high neutralization titers against D614G (ID50 = 5,489 - 16,760); these participants were boosted at least 4 months after the second dose. Shown are ID50 titers (A) and fold reduction in ID50 geometric mean titers (GMT) compared to D614G (B). Values below the limit of detection (ID50 = 20) were assigned a value of ID50 = 10. Bars extend from the 25^th^ to 75^th^ percentile. Horizontal lines and values above each bar in the left panels are GMT. Values above each bar in the right panels are the geometric mean fold change relative to D614G. Solid circles, two participants infected 5-6 months after second dose. Open circles, uninfected participants.

**Figure 2.**
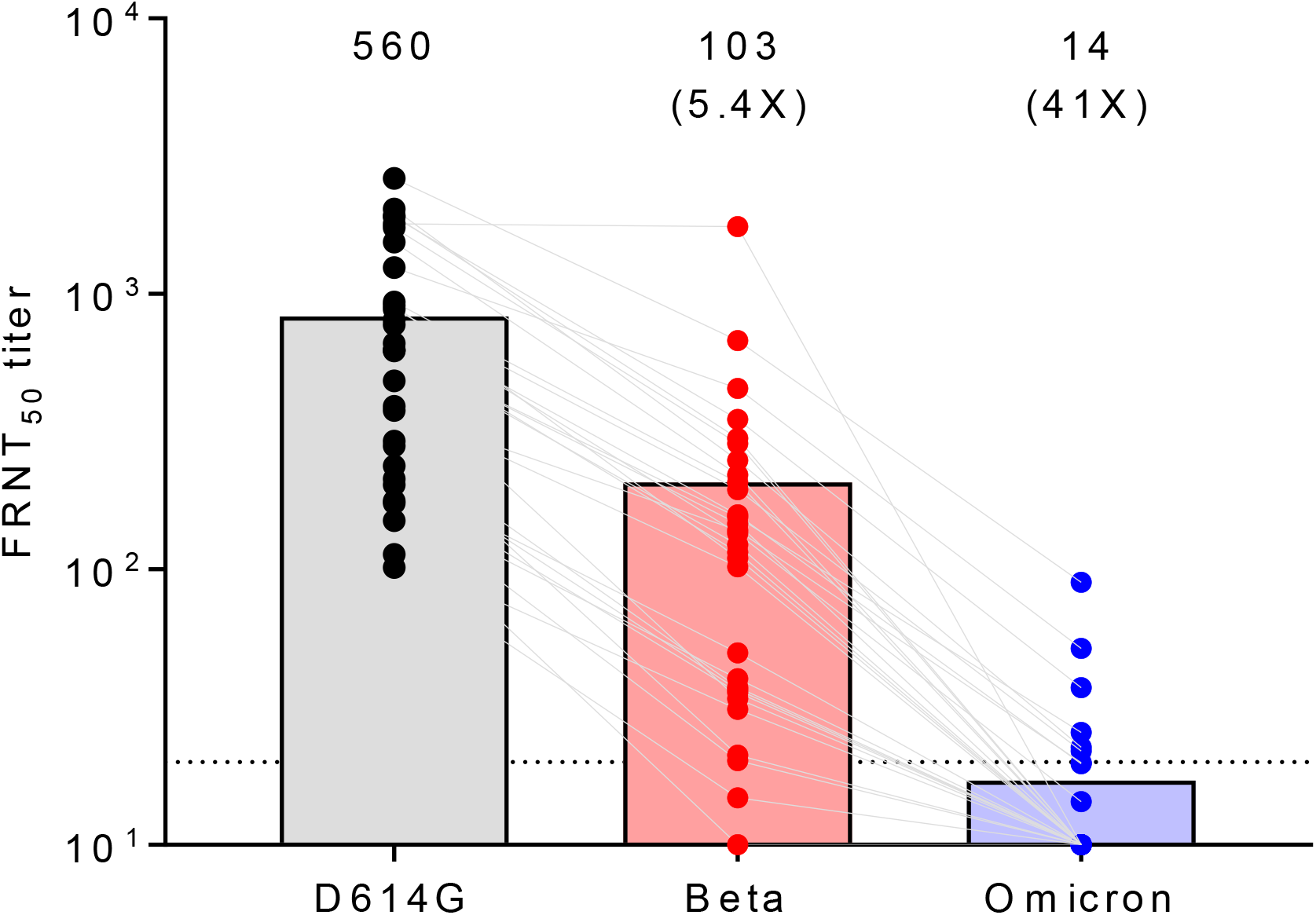
Neutralization of D614G, Beta and Omicron Live Viruses by Serum Samples Obtained from Moderna mRNA-1273 Vaccine Recipients. Serum samples obtained from the same 30 vaccine recipients 4 weeks after 2nd inoculation with mRNA-1273 in the phase 3 COVE study in Figure 1 were assayed in the live virus focus-reduction neutralization (FRNT) assay in the Emory laboratory. Shown are individual FRNT_50_ titers as solid dots, with GMT (values above each plot) and fold reduction in geometric mean titers (GMT) compared to D614G (values in parentheses). Values below the limit of detection (FRNT_50_ = 20) were assigned a value of FRNT_50_ = 10. Bars extend to the GMT.

In a separate analysis, sequential serum samples from the 7 participants who were vaccinated and boosted under EUA were tested against D614G, Beta and Omicron in the pseudovirus assay (VRC lab). These samples were obtained 2 weeks-post second dose (already assayed above), pre-boost, and 2 weeks post-boost. Two participants were infected 5-6 months after the second dose (prior to boost) at a time when Alpha and Delta were dominant variants in the USA. ID50 titers 2 weeks after the second dose were highest against D614G and lowest against Omicron, where titers against Omicron were 35.1-fold lower and titers against Beta were 8.9-fold lower than D614G (Fig 3). Titers for the 5 uninfected participants decreased substantially by the day of boosting and rose sharply 2 weeks post-boost to titers exceeding those after the second dose; this was true for all three variants. The increase in ID50 GMT post-boost compared with a post-second dose was 12.6-fold for Omicron and 6.7-fold for Beta in these 5 participants. In contrast to the dramatic reduction in titers seen after the second dose, the post-boost ID50 GMT against Omicron showed a more modest 6.5-fold reduction compared to D614G, and the post-boost titers against Beta showed a 3.4-fold reduction compared to D614G, for all 7 participants combined. Results for the two participants who were infected between dose 2 and the boost demonstrate that infection with an earlier variant can enhance the vaccine-response to all three variants, and that additional vaccine boosting can further increase this response (Fig 3).

**Figure 3.**
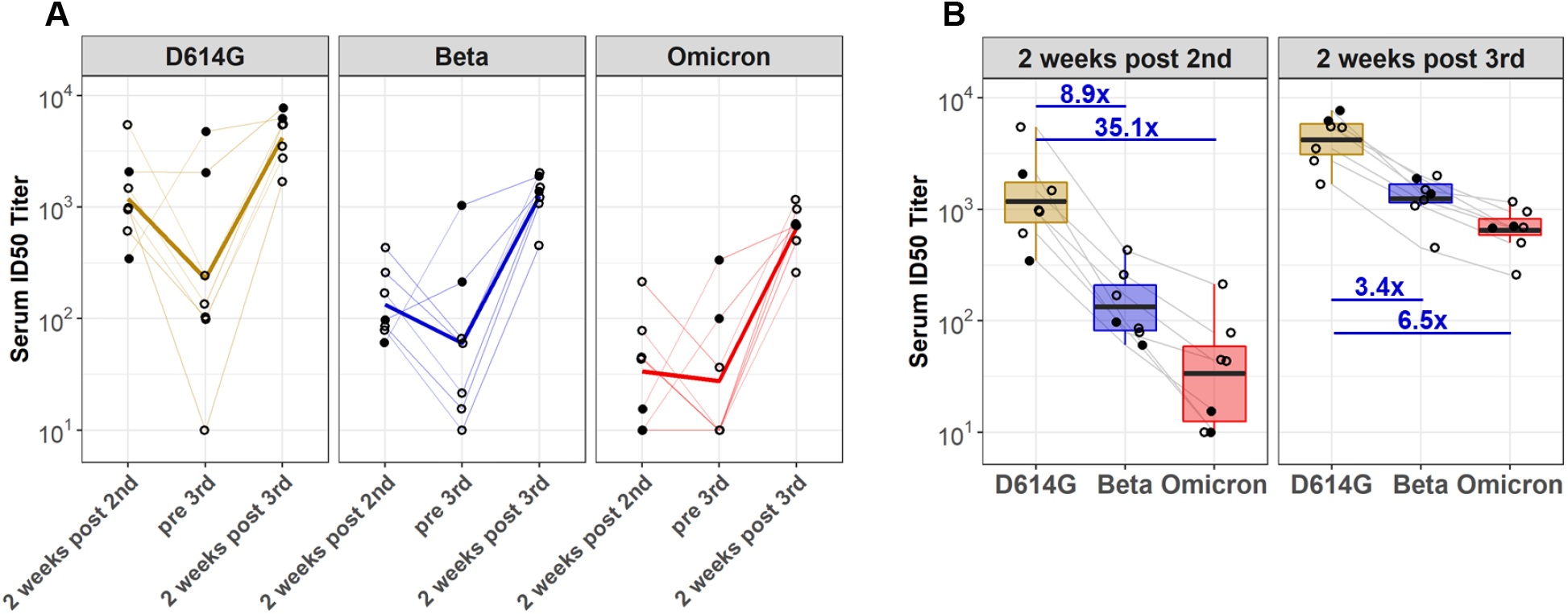
Longitudinal assessment of waning and recall neutralizing antibody responses in recipients of three doses of mRNA-1273. A. Shown are ID50 neutralization titers of serum samples from 7 recipients of three doses of mRNA-1273 assayed against D614G, Beta and Omicron. Samples were obtained 2 weeks after second dose, day of boost and 2 weeks post boost from participants who received the vaccine under EUA. Thin lines are individual samples. Heavy lines are geometric mean titers (GMTs) of all 7 values per time point. B. Aggregate ID50 titers for the two peak immune time points. Values below the limit of detection (ID50 = 20) were assigned a value of ID50 = 10. Bars extend from the 25^th^ to 75^th^ percentile. Horizontal lines within the bars, and values above each bar, are GMT. Also shown are the geometric mean fold reduction in ID50 compared to D614G. Solid circles, two participants who were infected 5-6 months after second dose. Open circles, uninfected participants.

Together, these results indicate that neutralizing titers to Omicron are 41-84 times lower than neutralization titers to D614G after 2 doses of mRNA-1273, which could lead to an increased risk of symptomatic breakthrough infections. However, a booster dose of mRNA-1273 increases Omicron neutralization titers and may substantially reduce this risk.

This study has several limitations: 1) relatively small sample sets that do not reflect the complete range of neutralization titers in diverse demographic groups of individuals, 2) limited longitudinal data, and 3) lack of data on the durability of Omicron neutralizing antibody responses post-boost. Additional work is in progress to address these limitations. The current study design permitted a preliminary assessment of the potential of Omicron to evade mRNA-1273-elicited neutralizing antibodies after 2 and 3 vaccine inoculations.

## Data Availability

All data produced in the present study are available upon reasonable request to the authors

## Acknowledgements

We are very grateful to the many study participants who donated specimens for this study. We thank Obrimpong Amoa-Awua, Man Chen, Wing-Pui Kong, Kwanyee Leung, Allen Mueller, Wei Shi, Eun Sung Yang, Tongqing Zhou, and the VRC200 Study Team of the VRC Clinical Trials program for technical assistance. This study received support from NIH 75N93019C00050 (DCM, XS), NIH U19 AI142790 (BK), NIH UM1AI148684 (LG), NIH UM1AI148575 (RLA), NIH UM1 AI148689 (KL), NIH/NIAID CEIRR under contract 75N93021C00017 (MSS), NIH 3U19AI057266-17S1 (MSS), NIH 1U54CA260563 (MSS), NIH P51OD011132 (MSS), Woodruff Health Sciences Center 2020 COVID-19 CURE Award (MSS), Moderna, Inc., and the Intramural Research Program of the Vaccine Research Center, NIAID, NIH (NADR, SDS, SOD, YZ, MRG, WPB, IG, MG, JEL, and JRM).

## References

1. Korber B, Fischer WM, Gnanakaran S, et al. Tracking changes in SARS-CoV-2 spike: evidence that D614G increases infectivity of the COVID-19 virus. Cell 2020; 182(4): 812–827.e19.

2. Baden LR, El Sahly HM, Essink B, et al. Efficacy and Safety of the mRNA-1273 SARS-CoV-2 Vaccine. N Engl J Med 2021; 384:403–416.

3. El Sahly, H.M., et al. Efficacy of the mRNA-1273 SARS-CoV-2 Vaccine at Completion of Blinded Phase. N Engl J Med 2021; 385:1774–1785.

4. Gilbert PB, Montefiori DC, McDermott AB, et al., Immune correlates analysis of the mRNA-1273 COVID-19 vaccine efficacy clinical trial. Science 2021; 10.1126/science.abm3425.

5. Pegu A, O’Connell SE, Schmidt SD, et al. Durability of mRNA-1273 vaccine–induced antibodies against SARS-CoV-2 variants. Science 2021; 373:1372–1377.

6. Bruxvoort KJ, Sy LS, Qian L, et al. Effectiveness of mRNA-1273 against Delta, Mu, and other emerging variants. medRxiv preprint doi: https://doi.org/10.1101/2021.09.29.21264199

7. Abu-Raddad LJ, Chemaitelly H, Butt AA. Effectiveness of the BNT162b2 Covid-19 vaccine against the B.1.1.7 and B.1.351 variants. N Engl J Med 2021; 385:187–189.

8. World Health Organization. 2021. Classification of Omicron (B.1.1.529): SARS-CoV-2Variant of Concern. https://www.who.int/news/item/26-11-2021-classification-of-omicron-(b.1.1.529)-sars-cov-2-variant-of-concern.

9. Hastie KM, Li H, Bedinger D, et al. Defining variant-resistant epitopes targeted by SARS-CoV-2 antibodies: A global consortium study. Science 10.1126/science.abh2315 (2021).

10. Edara VV, Pinsky BA, Suthat MS, et al. Infection and vaccine-induced neutralizing-antibody responses to the SARS-CoV-2 B.1.617 variants. N Engl J Med 2021; 385:664–666.

